# Comparison of local large language models for extraction of signs and symptoms data from electronic health records

**DOI:** 10.64898/2025.12.11.25341954

**Authors:** Isa Spiero, Merijn H. Rijk, Matthew A. Scheeres, Frans H. Rutten, Geert-Jan Geersing, Tamara N. Platteel, Karel G.M. Moons, Lotty Hooft, Johanna A.A. Damen, Roderick P. Venekamp, Artuur M. Leeuwenberg

**Affiliations:** Department of Epidemiology & Health Economics, Julius Center for Health Sciences and Primary Care, University Medical Center Utrecht, Utrecht University, Utrecht, the Netherlands; Department of General Practice & Nursing Science, Julius Center for Health Sciences and Primary Care, University Medical Center Utrecht, Utrecht University, Utrecht, the Netherlands; Cochrane Netherlands, University Medical Center Utrecht, Utrecht University, Utrecht, the Netherlands

## Abstract

Electronic health records (EHRs) provide a large source of data that can be used for research purposes. Extraction of information from unstructured clinical notes in EHRs can be automated by large language models (LLMs). Although LLMs are promising for this task, challenges remain in reliable application of LLMs to EHR, including the lack of development and validation for languages other than English. Here, we identified Dutch LLMs and compared their performance in a case study. We selected the MedRoBERTa.nl and RobBERT models based on local applicability, Dutch language compatibility, and model architecture. We evaluated their performance in a case study on the extraction of signs and symptoms from comprehensive Dutch primary care EHRs of patients with a lower respiratory tract infection. Using manually annotated clinical notes, models were trained as direct and prompt-based classifiers with varying amounts of training samples. Performance was expressed by precision, recall, and F1-score. The MedROBERTa.nl and RobBERT models showed good performance as direct classifiers, with a macro-averaged F1-score of 0.74 (range 0.56-0.87) and 0.69 (range 0.46-0.86) using 1600 training samples, respectively. The prompt-based classifiers performed worse with F1-scores of 0.08 (range 0.02-0.30) and 0.08 (range 0.02-0.22), respectively. In general, performance of the models was negatively affected by class imbalance and missingness of signs and symptoms. A minimum of 800 annotated training samples were required to obtain sufficient performance. The selected LLMs showed good performance as direct classifiers in extracting signs and symptoms from Dutch primary care EHRs. However, prompt-based models require performance improvement by further prompt engineering, and caution is warranted with imbalanced or partially missing EHR data.MedROBERTa.nl and RobBERT models, used as direct classifiers, can be considered for clinical research to extract information from clinical notes from Dutch primary care EHRs, potentially reducing manual annotation time and accelerating real-world research and evidence generation.

## Introduction

Electronic health records (EHRs) are increasingly used in clinical research as they form a valuable source of information on patients and health care use (1). Generally, EHRs contain structured data, such as disease codes and demographics, and unstructured data, such as clinical notes (2). While structured data can easily be retrieved, a large amount of valuable information is described by clinicians in unstructured data as narrative, free text in clinical notes. Manual screening of these clinical notes to retrieve information for research purposes is laborious and costly.

To facilitate extraction of unstructured data from clinical notes, natural language processing (NLP) techniques have been developed to automate information extraction for a wide range of EHR extraction tasks (1,3). Current growth in NLP techniques for extraction mainly involves the fast-paced development of large language models (LLMs), which are, in contrast to more simple NLP techniques, able to process texts in a more context-aware manner (4). These NLP techniques have already shown promising performance in extracting data from clinical notes in a wide range of studies and extraction targets such as specific clinical conditions (5), chronic diseases (6), symptoms (7), or social determinants of health (8).

Given the potential of LLMs to automatically extract unstructured EHR data, several areas of clinical research - such as prognostic prediction model studies - could benefit. Prognostic prediction modelling could be improved by applying LLMs to automatically extract large numbers of eligible patients, (candidate) predictors, and outcomes from EHRs. Using LLMs, study sample sizes can thus be increased and predictors reported or hidden in unstructured data can be included. This can subsequently improve the performance and the generalizability of the prediction model (9).

Still several challenges remain for reliable extraction of data from clinical notes in EHRs by LLMs. For example, EHR data are not developed for research purposes and therefore the information in clinical notes tends to contain abbreviations, grammatical errors, ambiguous language and incomplete or selective information, which could negatively impact the LLM’s extraction performance (10,11). In addition, training and validating LLMs for extraction often requires manually annotated clinical notes, which are costly to obtain and therefore only available in small amounts (10). Furthermore, the best performing LLMs are only available online and therefore not locally applicable in secured environments where EHR data are stored, which is crucial for the confidential data present in EHRs. And finally, LLMs are mostly developed for the English language (12), but the majority of patients worldwide receive care in other languages, creating a need for locally trained LLMs. Although development is growing for other languages (13), relatively little is known about the performance of LLMs for most languages including the application of LLMs to EHR notes in Dutch language.

We address these issues by exploring the performance of locally applicable LLMs for extraction of data from Dutch EHRs. We gain insight on the potential of LLMs for the extraction of EHR data to be used in, for example, prediction model research. First, we performed a literature search for locally applicable LLMs available for Dutch clinical texts. Subsequently, we applied the selected models to extract signs and symptoms from EHR data of adult patients presenting to primary care with a lower respiratory tract infection (LRTI), which are the target predictors for a prediction modeling case study described elsewhere (14). We additionally estimated the sample size of the training data needed for sufficient LLM performance.

## Materials & methods

### Case study: predicting 30-day hospitalization or mortality in primary care LRTI patients

We evaluated the performance of Dutch LLMs in a case study aiming to extract nine signs and symptoms from EHR data of adult LRTI patients presenting to their general practitioner (GP). These signs and symptoms, summarized in Table 1, are candidate predictors for a prognostic modelling study aiming to predict individual risk of 30-day mortality or hospitalization using EHR data of LRTI patients as described elsewhere (14). However, data on signs and symptoms are recorded in unstructured clinical notes and would require manual extraction for incorporation into such a prediction model. As this would be infeasible on large scales, LLMs could be implemented to automatically extract signs and symptoms from EHR clinical notes from large numbers of patients.

**Table 1.** Recording of signs and symptoms in clinical notes from EHR of 2,000 primary care LRTI patients, that are targets for extraction by the LLMs, from Rijk *et al*. (2024)

|  | Overall | Recorded as<br>'present' [overall], % | Recorded as<br>'absent' [overall], % | Not recorded,<br>% |
| --- | --- | --- | --- | --- |
| **Patient reported [Dutch]** |  |  |  |  |
| Cough [*Hoesten*] | 76.8 | 98.2 [75.4] | 1.8 [1.4] | 23.2 |
| Fever [*Koorts*] | 58.3 | 43.1 [25.1] | 56.9 [33.2] | 61.7 |
| Shortness of Breath [*Kortademigheid*] | 51.4 | 76.0 [39.1] | 24.0 [12.3] | 58.6 |
| Sputum [*Sputum*] | 26.2 | 89.5 [23.4] | 10.5 [2.8] | 73.8 |
| Chest pain [*Pijn op de borst*] | 14.7 | 72.4 [10.6] | 27.6 [4.1] | 85.3 |
| Chills [*Koude rillingen*] | 4.9 | 92.9 [4.6] | 7.1 [0.3] | 95.1 |
| **Patient or GP reported [Dutch]** |  |  |  |  |
| Confusion [*Verwardheid*] | 3.1 | 31.1 [1.0] | 68.9 [2.1] | 96.9 |
| **GP reported [Dutch]** |  |  |  |  |
| Crackles upon auscultation<br>[*Crepitaties*] | 84.3 | 27.5 [23.2] | 32.5 [61.1] | 15.7 |
| Ill appearance [*Zieke indruk*] | 30.3 | 35.3 [10.7] | 64.7 [19.6] | 69.7 |

GP, general practitioner; LRTI, lower respiratory tract infections; EHR, electronic health records; LLM, large language model.

### Data

We used data from a sample of 2,000 patients aged ≥40 years presenting to their GP with an LRTI between 2016 and 2019 in the region of Utrecht, The Netherlands (15). The data were derived from the comprehensive Julius General Practitioners’ Network (16), and contains both structured (e.g. demographics, medical history, medication use) and unstructured (e.g. descriptions of signs, symptoms, physical examination) data of the first GP consultation within an LRTI episode. Unstructured data was covered in ‘subjective’ (patient reported), ‘objective’ (GP reported), ‘evaluation’, and ‘plan’ text fields. We used the subjective and objective texts as our target for extraction, hereafter called ‘clinical notes’. Most clinical notes had between 5 and 100 tokens (words) with an average number of 63 tokens and a few large notes containing up to 240 tokens.

### Target variables

The target variables for extraction consisted of both patient and GP reported LRTI signs and symptoms (Table 1). The clinical notes were manually annotated for these signs and symptoms in a previous study by (15). Each sign or symptom was annotated for each patient as (*a*) ‘present’, (*b*) ‘absent’, or (*c*) ‘not

reported’. These annotations functioned as the reference standard for the LLM. The LRTI signs and symptoms show a strong variation in class imbalance/prevalence (i.e., some occur more rarely across patients than others) as well as missingness (i.e., some signs and symptoms are often not reported) (Table 1).

### Large Language Models

#### Possible approaches of LLMs

Generally, there are three approaches to implement LLMs for extraction of EHR data:

- <u>Training from scratch.</u> This approach involves completely building the LLM from the ground up, including designing the architecture of the LLM (in terms of layers, attention heads, embedding sizes, etc.). This allows for complete freedom in designing and altering the model which enables to tailor the model to the specific task at hand, at the cost of (generally) very high computational power needed as well as vast amounts of training data. In our setting, this computation was not feasible.
- <u>Using a pretrained model.</u> This is a simple approach in which a model is used that had previously been trained on either a general dataset or a domain-specific dataset. The pre-trained model is used as-is, and is directly applied to perform the desired task. Although this approach requires minimal effort and is the most straightforward, the model might not perform optimally as it is not optimized for the specific data and task at hand.
- <u>Fine-tuning a pretrained model.</u> This approach involves using a pretrained model which is then fine-tuned for the specific task at hand by further training the model with the available data. This allows the model to adapt to the desired task and does not require an extremely large dataset for training. For these reasons, extending a pretrained model is often the first choice for LLM-related research in domain-specific appliances, such as extraction from Dutch EHR data.

#### Possible implementations of LLMs for classification

For all three approaches described above, the LLMs can subsequently be used for classification tasks in several ways, of which two common methods are described below:

- <u>Direct classification.</u> In direct classification, a model learns from a set of training samples (here, clinical notes) to directly discriminate between a fixed set of predefined classes (here, ‘absent’, ‘present’, and ‘not reported’). After training, the model can be applied to other samples to predict to which class the new sample would belong (here, to predict for a clinical note if a given sign or symptom is ‘present’, ‘absent’, or ‘not reported’).
- <u>Prompt-based classification.</u> In prompt-based classification, the model is not directly trained for the classification task, but a so-called conversational LLM is used and prompted with a natural language prompt that asks the LLM using human language to classify the text that it is given. For these types of models, one of the following setup types can be applied:

- Zero-shot classification: The model classifies based solely on a task description. Example prompt: “Classify whether the patient in this text has fever or not: [text]”
- One-shot classification: The model classifies based on a task description and one labelled sample. Example prompt: ““While the patient coughs constantly, there is no sign of elevated temperature” => “No fever”. Classify whether the patient in this text has a fever or not: [text]”
- Few-shot or multishot classification: The model classifies based on a task description and two or more labelled samples. Example prompt: ““While the patient coughs constantly, there is no sign of elevated temperature” => “No fever”. “Fever was confirmed after measuring a temperature of 39.2 degrees Celsius” => “Fever”. Classify whether the patient in this text has a fever or not: [text]”

#### Our selection, approach, and implementation of LLMs

We first performed a search on Hugging Face (an open-source platform for AI and machine learning models) to find and select suitable pre-trained LLMs. Our selection criteria for LLMs included (1) locally applicability, (2) pre-trained, (3) a sufficient and locally computable number of parameters, (4) compatibility with Dutch language, and (5) preferably familiarized with medical texts (see the entire selection procedure in Appendix A). We selected two models fulfilling the criteria, MedRoBERTA.nl (17) and RobBERT (18) (see further model details in Appendix A).

We approached our task of extracting signs and symptoms from Dutch EHR data by fine-tuning the selected pre-trained LLMs. We implemented the models in two different ways for each model: as a *direct classifier* and as a *prompt-based classifier*, leading to the following four model configurations that were evaluated:

- direct classifier – MedRoBERTa.nl
- direct classifier – RobBERT
- prompt-based – MedRoBERTa.nl
- prompt-based – RobBERT

#### Implementing LLMs as direct classifiers

For direct classification, each of the models was trained using the manually annotated dataset to discriminate between the three classes of ‘present’, ‘absent’, and ‘not reported’. We trained each of the models for 5 epochs, a number which we empirically determined beforehand by assessing the training and evaluation loss of MedRoBERTa.nl after 10 epochs. To additionally evaluate the effect of training sample size on the performance of the direct classifier LLMs, we varied the number of training samples by 200, 400, 600, 800, 1000, 1200, 1400, and 1600. The performance of the models was evaluated by 5-fold cross-validation in which we used fixed folds across each experiment and kept the same 400 samples as the validation set.

### Implementing LLMs as prompt-based classifiers

Before we could use the models as prompt-based classifiers, we had to convert them to process Dutch prompts and apply those to Dutch EHR data. The following steps were taken to convert the original models to prompt-based classifiers:

1. First, we transformed the models to be able to also *generate* text, as the prompt-based classifiers predict labels through next token prediction. Therefore, we combined the encoder structure of the original models with the base multilingual BERT decoder using a Hugging Face Transformers EncoderDecoderModel^2^ architecture.
2. Second, all model parameters (including the decoder parameters) were fine-tuned for 3 epochs using a dataset of over 100,000 patient-doctor conversations (the HealthCareMagic-100k dataset^1^ from (19)) which was translated beforehand to Dutch using the Google Translate API.
3. After fine-tuning, a prompt could be used in which the multiclass prediction task is instructed to the model by providing a sample and an instruction to classify based on some provided samples.

The architecture of our prompts is shown in Box 1 in which the amount of input samples in the prompt for these models would be varied between 1, 2 or 3 samples.

#### Box 1. The prompt (for this article translated to English) that was used for the prompt-based models to extract the signs and symptoms from the Dutch EHRs. The original Dutch prompts are presented in Appendix C Box C1

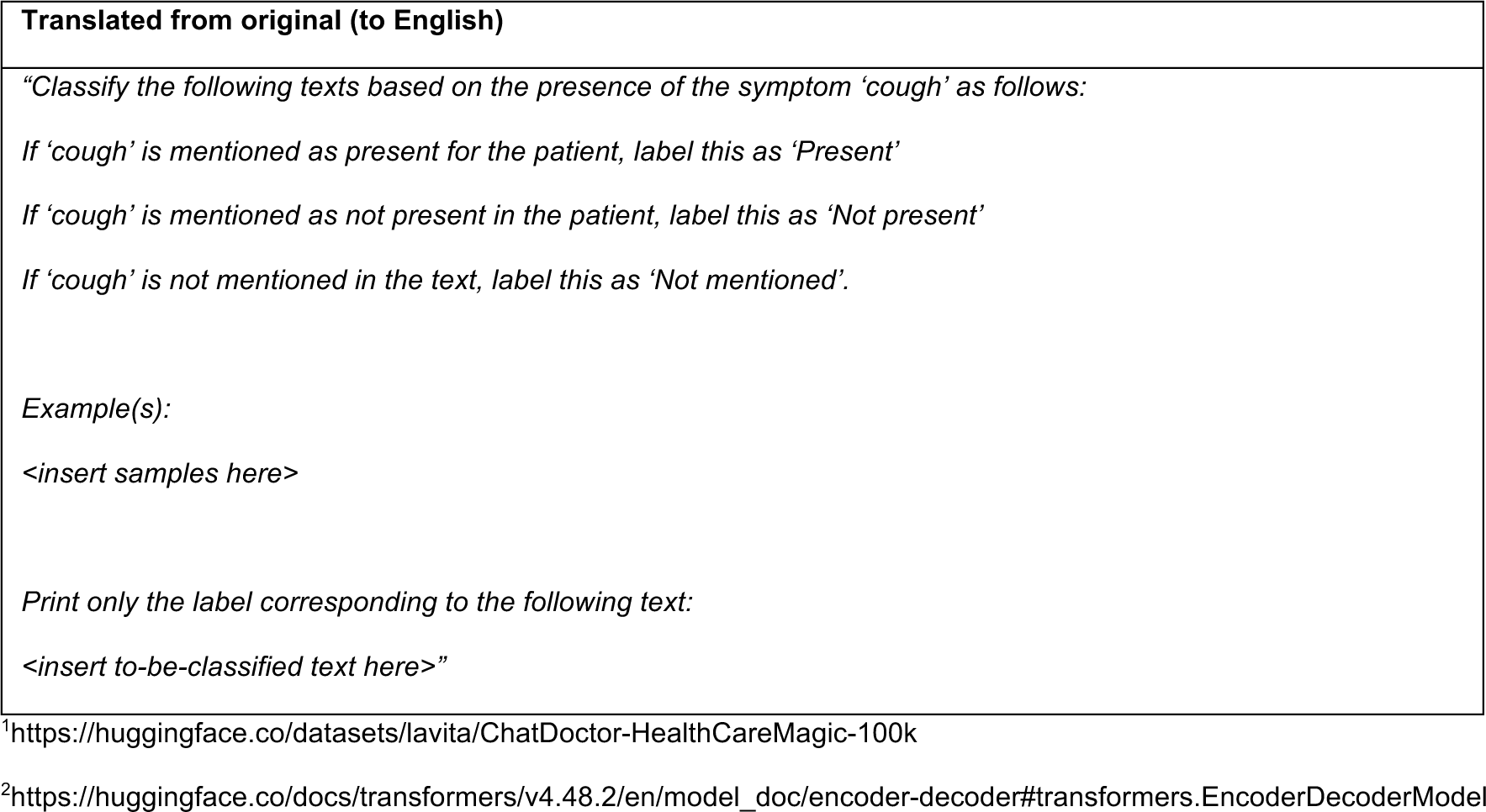

| Translated from original (to English) |
| --- |
| *"Classify the following texts based on the presence of the symptom 'cough' as follows:*<br>*If 'cough' is mentioned as present for the patient, label this as 'Present'*<br>*If 'cough' is mentioned as not present in the patient, label this as 'Not present'*<br>*If 'cough' is not mentioned in the text, label this as 'Not mentioned'.*<br>*Example(s):*<br>*<insert samples here>*<br>*Print only the label corresponding to the following text:*<br>*<insert to-be-classified text here>"* |

<sup>1</sup>https://huggingface.co/datasets/lavita/ChatDoctor-HealthCareMagic-100k
<sup>2</sup>https://huggingface.co/docs/transformers/v4.48.2/en/model_doc/encoder-decoder#transformers.EncoderDecoderModel

### Performance evaluation

In total, separate LLMs were developed and evaluated for each of the 9 target signs/symptoms and for each of the training sample sizes, leading to a total of 2 (direct classifiers) * 9 (signs and symptoms) * 8 (training sample sizes) + 2 (prompt-based classifiers) * 9 (signs and symptoms) * 3 (training sample sizes) = 198 models.

To evaluate and compare the performance of the LLMs, we computed the following metrics and averaged these across the 5-fold cross-validations:

- Precision (P): Precision is the ratio of true positive predictions to the total amount of positive predictions (a combination of true positives (TP) and false positives (FP)), i.e. the relative amount of correctly predicted positive samples, and is calculated by 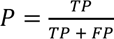.
- Recall (R): Recall quantifies how well the model is able to identify positive cases, i.e. all cases where a symptom is present in the text. Recall is calculated using the formula 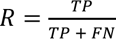.
- F1-score (F1): The F1-score provides a harmonic mean of precision and recall and functions as a balanced measure of both metrics. The F1-score is calculated using the formula *F*1 = 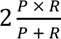.

The performance metrics were computed per-class and as macro-average across classes.

### Implementation details

The experiments were conducted in Python v3.11.5 on a virtual computer with 32 GB of RAM, an Intel(R) Xeon(R) Platinum 8272CL CPU, using an NVIDIA T4 GPU.

## Results

### Model comparison: MedRoBERTa.nl versus RobBERT

#### Direct classification

As direct classifiers, the MedRoBERTa.nl and RobBERT models performed similarly in classifying the presence of LRTI signs and symptoms from the Dutch clinical notes (Fig 1a). For the MedRoBERTa.nl model at the largest number of 1600 training samples used, the obtained recall in direct classification was on average 0.72 (range 0.54 - 0.86), the precision 0.76 (range 0.60 - 0.90), and the F1-score 0.74 (range 0.56 - 0.87) across the signs and symptoms. For the RobBERT model at the largest number of 1600 training samples used, the obtained recall in direct classification was on average 0.69 (range 0.45 - 0.84), the precision 0.71 (range 0.48 - 0.89), and the F1-score 0.69 (range 0.46 - 0.86) across the signs and symptoms (Appendix B Table A3).

**Figure 1.**
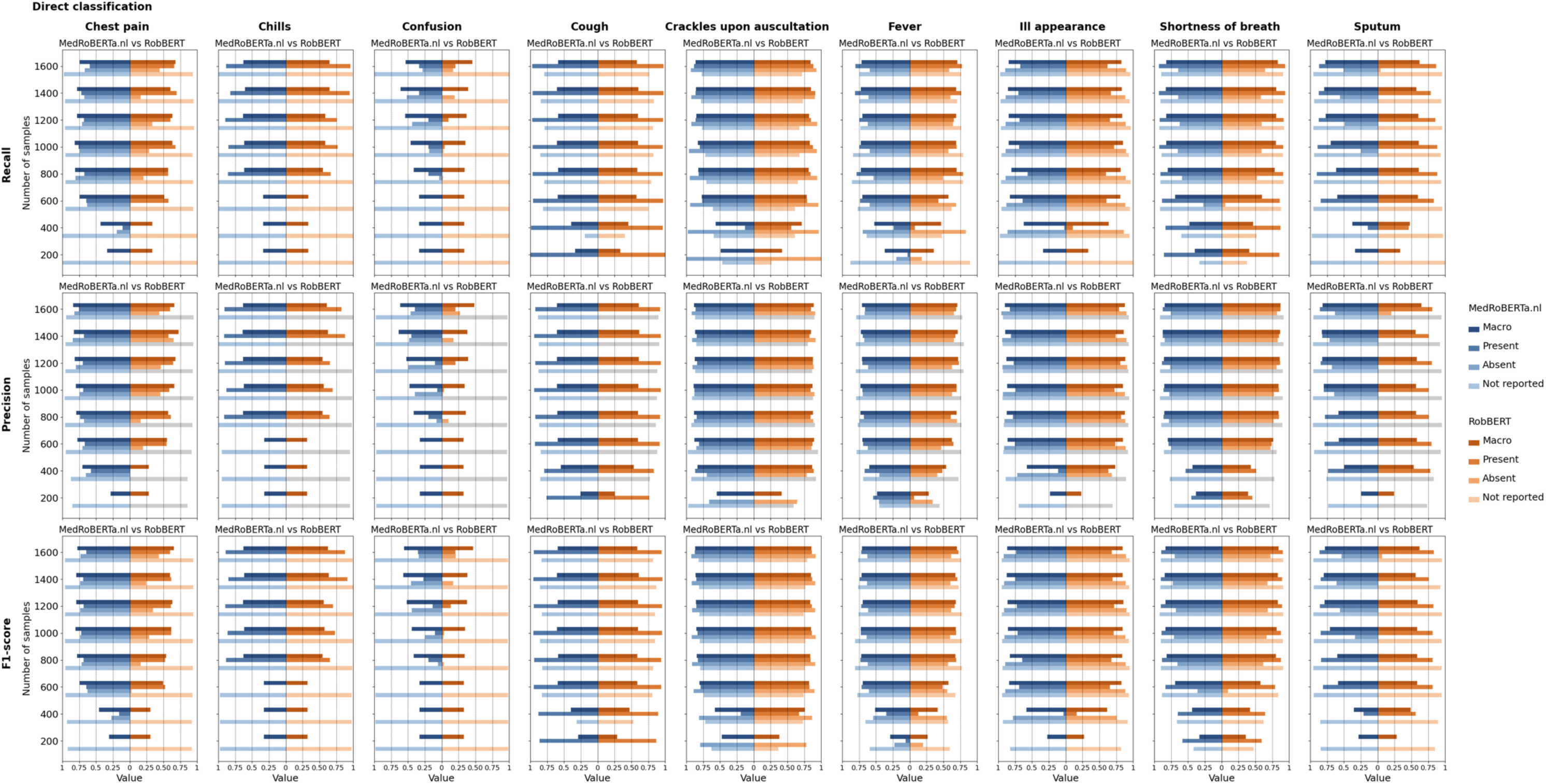
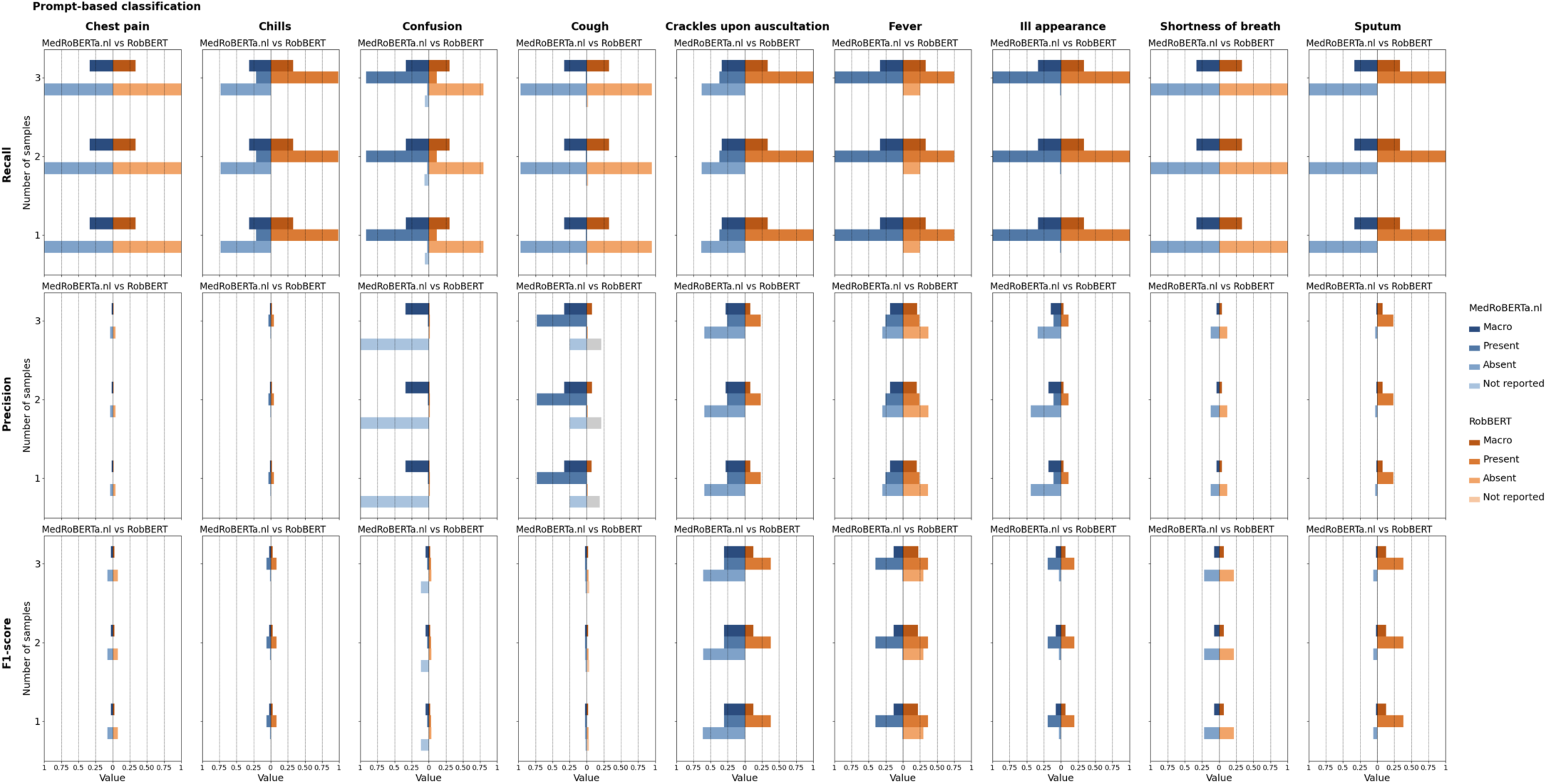
| The performance of the MedRoBERTa.nl and RobBERT models in classifying signs and symptoms related to lower respiratory tract infection based on clinical notes in Dutch primary care electronic health records. The performance is expressed in terms of the macro-averaged and per-class (’present’, ‘absent’, or ‘not reported’) recall, precision, and F1-score. The two models were implemented as (a) direct classifier trained on 200, 400, 600, 800, 1000, 1200, 1400 or 1600 training samples and (b) prompt-based classifier trained with 1, 2, or 3 samples.

#### Prompt-based classification

When the MedRoBERTa.nl and RobBERT models were implemented as prompt-based classifiers, the performance was significantly lower compared to direct classification, and less similar between the two models (Fig 1b). At the largest number of 3 samples used, for the MedRoBERTa.nl model the obtained recall in prompt-based classification was on average 0.33 (range 0.32 - 0.34), precision 0.15 (range 0.01 - 0.34), and F1-score 0.08 (range 0.02 - 0.30), while for the RobBERT model the obtained recall in prompt-based was on average 0.33 (range 0.31 - 0.33), precision 0.06 (range 0.01 - 0.21), and F1-score 0.08 (range 0.02 - 0.22) across the signs and symptoms (Appendix B Table A4). For most signs and symptoms, the prompt-based models conflated to predict one outcome for all data points. Hence, the recall values of 1.0 for one of the outcomes and close to 0 for the others, indicating poor model performance (Fig 1b).

### Effect of sign and symptom reporting and class imbalance

#### Direct classification

The level of reporting of a sign or symptom across the clinical notes significantly impacted the MedRoBERTa.nl and RobBERT models as direct classifiers in their performance. For example, in classifying two of the symptoms that were rarely reported, ‘Chills’ (reported in 4.9%) and ‘Confusion’ (reported in 3.1%), the models performed worse compared to more commonly reported signs and symptoms (Fig 1a). Moreover, the degree of class imbalance (i.e., the balance between ‘present’ and ‘absent’) also seemed to affect the performance of the models. For example, the extraction of the well-balanced symptoms of ‘Fever’ (43.1% ‘present’ if recorded), ‘Ill appearance’ (35.3% ‘present’ if recorded), and ‘Crackles upon auscultation’ (27.5% ‘present’ if recorded) showed the highest performance with F1-scores up to 0.87 compared to less balanced symptoms (Fig 1a).

#### Prompt-based classification

In the prompt-based classification, the RobBERT model performed better at classifying ‘Fever’ while the MedRoBERTa.nl model performed better at classifying ‘Crackles upon auscultation’ (Fig 1b). For the other signs and symptoms, there were no marked differences between the two models, but there were some small overall model-independent differences between the signs and symptoms. However, those differences were not related to the level of recording of those signs and symptoms in the clinical notes or to the balance in positive and negative recordings (Fig 1b).

### Effect of training sample size

#### Direct classification

In direct classification the performance of both models generally increased with an increasing number of training samples (Fig 1a). Recall ranged from 0.33-0.49 (MedRoBERTa.nl) and 0.26-0.38 (RobBERT) with 200 training samples to 0.54-0.86 (MedRoBERTa.nl) and 0.45-0.84 (RobBERT) with 1600 training samples, precision ranged from 0.23-55 (MedRoBERTa.nl) 0.23-0.41 (RobBERT) with 200 training samples to 0.60-0.90 (MedRoBERTa.nl) and 0.48-0.89 (RobBERT) with 1600 training samples, and F1-score ranged from 0.27-0.47 (MedRoBERTa.nl) and 0.26-0.38 (RobBERT) with 200 training samples to 0.56-0.87 (MedRoBERTa.nl) and 0.46-0.86 (RobBERT) with 1600 training samples (Appendix B Table A3). For both models, the classification performance for some symptoms, such as ‘Chills’ and ‘Confusion’, was generally low and, especially for ‘Confusion’, did not increase with more training samples. Generally, models’ performance reached a plateau by a minimum of 800 training samples (Fig 1a).

#### Prompt-based classification

When implementing the MedRoBERTa.nl and RobBERT models as prompt-based classifiers, the number of samples in the prompts (i.e., 1, 2, or 3) did not affect the performance of the models (Fig 1b). This was similar for both models and across all signs and symptoms.

## Discussion

We evaluated the performance of two LLMs for the Dutch language, RobBERT and MedRoBERTa.nl, for extracting signs and symptoms of LRTI patients from Dutch EHR data. Overall, the models performed similarly, but variation was observed across implementation methods (direct classification outperformed prompt-based classification), across signs and symptoms that were extracted (higher performance for signs or symptoms with limited missingness and class-imbalance), and across the sample size of the training data. Both models showed good performance when implemented as direct classifiers and trained with sufficient data (at least 800 samples) with F1-scores reaching up to 0.87.

The MedRoBERTa.nl and RobBERT models showed similar overall performance although we anticipated that the MedRoBERTa.nl model – which was pre-trained on domain-specific medical data – would outperform RobBERT for the domain-specific task of this study (20). However, the RobBERT model was originally trained on larger collections of data (18), which may explain why this model – not specifically trained on medical data – performed similarly as MedRoBERTa.nl. This may imply that LLMs pre-trained with large amounts of general domain data may equally suffice in extracting medical information from EHR data compared to LLMs trained with smaller amounts of medical domain data. Contrary, in the study by (21) MedRoBERTa outperformed all other tested models including RobBERT in classifying reports, suggesting that the performance of models is also context or task dependent.

For both LLMs, implementation as prompt-based classifier showed significantly worse performance compared to the direct classifier method. Although conceptually interesting as they only require an instruction and thus avoid the need for costly manually annotated data, the performance of the prompt-based classifiers in this study was low. This is possibly due to the limited size of the prompt and the numbers and/or content of the examples in the prompt, or to the fact that these models were not trained as an encoder-decoder model from the ground up. In a similar clinical note classification task, incorporation of a reasoning step as a part of the prompt, in which the model is explicitly asked to output its stepwise reasoning, did increase performance (22). Further engineering to optimize prompts may thus be needed to improve the performance of the prompt-based classifiers (23,24).

Substantial difference in model performance was also observed across the nine signs and symptoms that were extracted by the LLMs. For some, such as ‘crackles upon auscultation’, ‘ill appearance’, and ‘shortness of breath’, the direct classifiers showed promising performance, while for others the LLMs performed significantly worse. The differences in performance across signs and symptoms were likely related to class imbalance and missingness (non-reporting), with worse performance for more imbalanced and/or less often reported signs and symptoms. This, however, will remain a problem inherent to EHR data, despite mitigation methods (25). In addition, the variation in performance across the signs and symptoms may also be partly explained by the amount of variation in descriptions of signs or symptoms that clinicians use. For example, ‘ill appearance’ and ‘confusion’ may be reported in a wider variety of descriptions than other signs and symptoms, although we did not further quantify this nor explored the potential effect on model performance.

As expected, for most signs and symptoms the performance increased with increasing sample sizes of training data for direct classifiers, while the prompt-based classifiers showed no difference in performance between the number of training samples. We found that generally a minimum of 800 training samples is needed to obtain good model performance, with F1-scores up to 0.87 for some of the signs or symptoms for the MedRoBERTa.nl model. This implies that, with manual data annotation being laborious and resource-consuming, a relatively limited amount of training samples may suffice to train LLMs for an EHR extraction task.

### Strengths and limitations

This study is one of the first to evaluate and compare the performance of LLM in extracting signs and symptoms from Dutch unstructured EHR data. Further strengths of this study include the thorough search for locally applicable Dutch LLMs, the comparison between direct and prompt-based classifying models, and the large amount of manually annotated training samples which allowed us to determine the minimally required sample size of training data. Nevertheless, some limitations should be considered when interpreting these results. First, although we performed a careful selection with reasonable selection criteria, we only tested two models in our study. The performance of other models, especially non-local models that could not be implemented in our EHR setting, could not be determined. Also, larger models, which could not be run in our environment, hold promise to significantly improve performance (26,27). Second, we did not extensively engineer our prompts, which likely explains why the prompt-based classifiers underperformed. We did not further elaborate on optimal prompt engineering in this study as our focus was on the exploration and comparison of currently available models for Dutch EHR data extraction. Lastly, given the large variability in performance between the signs and symptoms, the insights of the current study may be highly context specific, and the performance of the models may not be generalizable to other data or clinical settings.

## Conclusion

After a thorough search for currently available LLMs for the extraction of signs and symptoms from Dutch EHR data, we selected the MedRoBERTa.nl and RobBERT models and compared their performance in a case study. Both models demonstrated good performance when implemented as direct classifiers and are therefore promising for the extraction of signs and symptoms from Dutch unstructured EHR data, while prompt-based classifiers consistently showed worse performance in our analyses. This study also found that a minimal training sample size of 800 samples was generally sufficient to train the models as direct classifiers for extraction of most signs and symptoms. These findings support the feasibility of using LLMs for scalable and automated symptom extraction in Dutch clinical research settings. However, model performance varied substantially across signs and symptoms, with better results for clinically structured and consistently reported findings, and lower performance for more sparsely documented symptoms. This highlights the need for clinical awareness when interpreting model outputs and suggests that symptom-specific strategies may be required. In the context of prediction research, future studies should assess whether including signs and symptoms that are automatically extracted from Dutch EHR data by LLMs leads to improvement of clinical prediction model performance and, ultimately, improves prediction of individual patient outcomes’ risks. Moreover, further development in prompt engineering may enhance performance and thereby reduce the need for costly manual annotations of clinical notes. These steps are essential to move from technical feasibility to meaningful clinical impact.

## Data Availability

Data cannot be shared publicly because data access is restricted, and has been granted under license of the current study. Data are therefore only available from the authors upon reasonable request and after formal permission of the JGPN. The codes corresponding to this study can be found at https://github.com/isa-sp/Local-LLMcomparison-LRTI

## Acknowledgements

The authors kindly acknowledge Marloes M. van Beurden for contributing to this study by extracting the relevant data from the Julius General Practitioners’ Network database.

