## Appendix for "Comparison of local large language models for extraction of signs and symptoms data from electronic health records"

### A. SELECTION OF LARGE LANGUAGE MODELS

#### Selection criteria

We defined a set of selection criteria that indicate the suitability of a large language model (LLM) for the purpose of our study which targets the extraction of data from Dutch electronic health records (EHRs):

- *Local applicability*: Since clinical notes and EHRs in general contain a multitude of privacy sensitive data, it is mandatory that the model is able to be completely implemented on local hardware, as no EHR data is allowed to be transmitted from the environment it is stored on. For this reason, only open-source, locally applicable LLMs could be selected.
- *Availability of a pre-trained model*: The model must be pre-trained, as we do not have the computational power and amount of data to train a model from scratch. We do however want to be able to further fine-tune our models on our own data and convert them into prompt-based classifiers as well.
- *Parameter count of a model*: The number of parameters in a model represents the size and complexity of a model and thus correlates with the computational power required to run it. As computational power is a limiting factor due to local application of the model, we needed to balance between having enough parameters for sufficient LLM performance and the ability to run the LLM run on a virtual desktop.
- *Dutch language competency*: It is vital for the model to be able to handle Dutch language data, either through optimization for the Dutch language or being completely trained on a Dutch dataset.
- *Medical text familiarity*: Models trained or fine-tuned on medical texts are more likely to be able to correctly process our domain-specific knowledge and jargon and are thus preferred.

**Model search**

To retrieve a list of relevant models, a literature search was performed and combined with a direct search for models on the Hugging Face website. These websites provide comprehensive filtering options, enabling us to find models that can be run offline rather easily. To find the models via HuggingFace, the following filters were applied during a search performed on 18 October 2023:

- _Tasks_: Text Classification, Zero-Shot Classification, Conversational, Text Generation, Fill-Mask
- _Libraries_: PyTorch, TensorFlow, Transformers, Sentence Transformers, Adapter Transformers,
- _Datasets_: *Not specified*
- _Languages_: Dutch
- _Licences_: *Not specified*
- _Other_: *Not specified*

In the context of this research, we focused our search of models on LLMs that show promise in classification tasks. The final list of potential models for those tasks and their relevant features is presented in Table A1.

**Table A1** | The large language models that were retrieved by our search including the characteristics on suitability for extraction of information from Dutch electronic health records.

| Model | Base Model | Number of parameters | Prompt-based model available | Local | Pretrained | Dutch | Medical | Open-source | Relevant paper | Model link | Suitable |
|---|---|---|---|---|---|---|---|---|---|---|---|
| *BERTje* | BERT | 109M | No | Yes | Yes | Yes | No | Yes | Link | GH \| HF | Yes |
| *GPT-2* (recycled for Dutch) | GPT-2 | 129M or 369M | No | Yes | Yes | Yes | No | Yes | Link | HF small, medium | Yes |
| *GPT-2 XL* | GPT-2 | 1.5B | No | Yes | Yes | Multilingual | No | Yes | Link | HF | Yes |
| *DistilBERT-nl* | BERT | 69M | No | Yes | Yes | Yes | No | Yes | Link | HF | Yes |
| *GPT-3.5/4* | GPT-3.5/4 | 154B / 1.76T | Yes | No | Yes | Multilingual | No | No | Link3.5 / Link4 | NA | No |
| *Legal BERT* | BERT | 295M | No | Yes | Yes | Yes | No | Yes | Link | HF | No |
| *medRoBERTa.nl* | BERT | 117M | Yes | Yes | No (possibly via contact) | Yes | Yes | Yes | Link | GH | Yes |
| *Google Bard* | Lambda | 137B | Yes | No | Yes | Multilingual | No | No | NA | NA | No |
| *LLaMa* | LLaMa | 65B | Yes | Yes | Yes | Multilingual | No | No | Link | HF | Yes |
| *LLaMa 2* | LLaMa2 | 7B to 70B | Yes | Yes | Yes | Multilingual | No | No | Link | HF | Yes |
| *ChatDoctor* | LLaMa-7B | 7B | Yes | Yes | No | Multilingual | Yes | No | Link | GH | Yes |
| *RoBERTa* | BERT | 355M | Yes | Yes | Yes | Multilingual | No | Yes | Link | HF | Yes |
| *RobBERT-2023* | BERT | 117M | Yes | Yes | Yes | Yes | No | Yes | Link | HF | Yes |
| *DialoGPT* | None | 117M, 345M or 762M | Yes | Yes | Yes | Multilingual | No | Yes | Link | GH | Yes, but not used |

**Final selection**

After our search for available models, two models were selected as most suitable for our task (see computation details of each model in Table A2):

- *MedRoBERTa.nl* [1]
    - Description: A Dutch medical model created by researchers at Vrije Universiteit Amsterdam, who used 13GB of text data from Dutch hospital notes to train an altered version of the RoBERTa model. This model has shown to perform better on this data in odd-one-out tasks when compared to general Dutch LLMs and is thus likely to perform well in our research.
    - Motivation: Of all chosen models, MedRoBERTa.nl is the only one directly trained on Dutch medical data. Consequently, this model is expected to perform very well on the dataset. One limitation is that the parameter count of the model is rather low by modern standards, which, while being a positive factor in terms of computational efficiency, might reduce its performance.
- *RobBERT* [2]
    - Description: RobBERT is a model built upon the foundation of BERT, serving as the core architecture. BERT was released as a multilingual model, and RobBERT was fine-tuned as a language-specific model, as research points out that doing so generally results in higher performance.
    - Motivation: RobBERT's strong performance on various NLP tasks and proven ability to handle Dutch text make it a suitable candidate for our investigation.

**Table A2** | The number of parameters of the selected MedRoBERTa.nl and RobBERT models. Both models were implemented as direct classifier and as prompt-based classifier. Prompt-based classifiers had a larger number of parameters compared to the direct classifiers due to extra layers being added by the EncoderDecoder model (with the base multilingual BERT model as decoder).

| Classifier | Model | Number of parameters |
|---|---|---|
| **Direct** | *MedRoBERTa.nl* | 125,980,419 |
| | *RobBERT* | 116,764,419 |
| **Prompt-based** | *MedRoBERTa.nl* | 263,859,258 |
| | *RobBERT* | 254,643,258 |

[1] Verkijk, S., & Vossen, P. (2021). MedRoBERTa. nl: a language model for Dutch electronic health records. In *Computational Linguistics in the Netherlands* (Vol. 11, pp. 141-159).

[2] Delobelle, P., Winters, T., & Berendt, B. (2020). Robbert: a dutch roberta-based language model. *arXiv preprint arXiv:2001.06286*.

### B. Performance metrics of the LLMs in extracting signs and symptoms from Dutch EHRs

**Table A3** | The performance of the MedRoBERTa.nl and RobBERT models as direct classifier in terms of recall, precision, and F1-score for different training sample sizes. The models were trained to extract a total of nine signs and symptoms related to lower respiratory tract infection from Dutch primary care electronic health records.

| | | | | | | Per class metrics | | | | | | | | |
|---|---|---|---|---|---|---|---|---|---|---|---|---|---|---|
| | | | Macro-averaged metrics | | | 'Present' | | | 'Absent' | | | 'Not reported' | | |
| **Model** | **Sign or symptom** | **Number of samples** | **Precision** | **Recall** | **F1-score** | **Precision** | **Recall** | **F1-score** | **Precision** | **Recall** | **F1-score** | **Precision** | **Recall** | **F1-score** |
| MedRoBERTa.nl | Chest pain | 200 | 0,28 | 0,33 | 0,31 | 0,00 | 0,00 | 0,00 | 0,00 | 0,00 | 0,00 | 0,85 | 1,00 | 0,92 |
| MedRoBERTa.nl | Chest pain | 400 | 0,70 | 0,43 | 0,45 | 0,58 | 0,10 | 0,16 | 0,65 | 0,19 | 0,27 | 0,87 | 0,99 | 0,93 |
| MedRoBERTa.nl | Chest pain | 600 | 0,77 | 0,74 | 0,74 | 0,67 | 0,65 | 0,65 | 0,70 | 0,63 | 0,63 | 0,95 | 0,95 | 0,95 |
| MedRoBERTa.nl | Chest pain | 800 | 0,79 | 0,81 | 0,78 | 0,74 | 0,68 | 0,68 | 0,68 | 0,80 | 0,72 | 0,96 | 0,95 | 0,95 |
| MedRoBERTa.nl | Chest pain | 1000 | 0,80 | 0,82 | 0,80 | 0,69 | 0,76 | 0,72 | 0,74 | 0,74 | 0,73 | 0,96 | 0,95 | 0,96 |
| MedRoBERTa.nl | Chest pain | 1200 | 0,82 | 0,78 | 0,79 | 0,70 | 0,68 | 0,68 | 0,80 | 0,71 | 0,74 | 0,96 | 0,96 | 0,96 |
| MedRoBERTa.nl | Chest pain | 1400 | 0,83 | 0,78 | 0,80 | 0,68 | 0,72 | 0,69 | 0,84 | 0,67 | 0,74 | 0,96 | 0,96 | 0,96 |
| MedRoBERTa.nl | Chest pain | 1600 | 0,84 | 0,74 | 0,78 | 0,75 | 0,59 | 0,65 | 0,82 | 0,67 | 0,73 | 0,94 | 0,97 | 0,96 |
| MedRoBERTa.nl | Chills | 200 | 0,32 | 0,33 | 0,32 | 0,00 | 0,00 | 0,00 | 0,00 | 0,00 | 0,00 | 0,95 | 1,00 | 0,97 |
| MedRoBERTa.nl | Chills | 400 | 0,32 | 0,33 | 0,32 | 0,00 | 0,00 | 0,00 | 0,00 | 0,00 | 0,00 | 0,95 | 1,00 | 0,97 |
| MedRoBERTa.nl | Chills | 600 | 0,32 | 0,33 | 0,32 | 0,00 | 0,00 | 0,00 | 0,00 | 0,00 | 0,00 | 0,95 | 1,00 | 0,97 |
| MedRoBERTa.nl | Chills | 800 | 0,64 | 0,62 | 0,63 | 0,91 | 0,86 | 0,88 | 0,00 | 0,00 | 0,00 | 0,99 | 1,00 | 1,00 |
| MedRoBERTa.nl | Chills | 1000 | 0,62 | 0,62 | 0,62 | 0,88 | 0,85 | 0,86 | 0,00 | 0,00 | 0,00 | 0,99 | 1,00 | 0,99 |
| MedRoBERTa.nl | Chills | 1200 | 0,63 | 0,63 | 0,63 | 0,90 | 0,90 | 0,90 | 0,00 | 0,00 | 0,00 | 0,99 | 1,00 | 1,00 |
| MedRoBERTa.nl | Chills | 1400 | 0,64 | 0,61 | 0,62 | 0,92 | 0,82 | 0,85 | 0,00 | 0,00 | 0,00 | 0,99 | 1,00 | 1,00 |
| MedRoBERTa.nl | Chills | 1600 | 0,63 | 0,63 | 0,63 | 0,91 | 0,89 | 0,89 | 0,00 | 0,00 | 0,00 | 0,99 | 1,00 | 1,00 |
| MedRoBERTa.nl | Confusion | 200 | 0,32 | 0,33 | 0,33 | 0,00 | 0,00 | 0,00 | 0,00 | 0,00 | 0,00 | 0,97 | 1,00 | 0,98 |
| MedRoBERTa.nl | Confusion | 400 | 0,32 | 0,33 | 0,33 | 0,00 | 0,00 | 0,00 | 0,00 | 0,00 | 0,00 | 0,97 | 1,00 | 0,98 |
| MedRoBERTa.nl | Confusion | 600 | 0,32 | 0,33 | 0,33 | 0,00 | 0,00 | 0,00 | 0,00 | 0,00 | 0,00 | 0,97 | 1,00 | 0,98 |
| MedRoBERTa.nl | Confusion | 800 | 0,42 | 0,41 | 0,41 | 0,20 | 0,20 | 0,20 | 0,08 | 0,04 | 0,05 | 0,97 | 1,00 | 0,99 |
| MedRoBERTa.nl | Confusion | 1000 | 0,48 | 0,46 | 0,44 | 0,07 | 0,20 | 0,10 | 0,40 | 0,19 | 0,25 | 0,98 | 1,00 | 0,99 |
| MedRoBERTa.nl | Confusion | 1200 | 0,52 | 0,54 | 0,52 | 0,10 | 0,20 | 0,13 | 0,49 | 0,44 | 0,44 | 0,98 | 0,99 | 0,99 |

| | | | | | | Per class metrics | | | | | | | | |
|---|---|---|---|---|---|---|---|---|---|---|---|---|---|---|
| | | | Macro-averaged metrics | | | ‘Present’ | | | ‘Absent’ | | | ‘Not reported’ | | |
| **Model** | **Sign or symptom** | **Number of samples** | **Precision** | **Recall** | **F1-score** | **Precision** | **Recall** | **F1-score** | **Precision** | **Recall** | **F1-score** | **Precision** | **Recall** | **F1-score** |
| MedRoBERTa.nl | Confusion | 1400 | 0,64 | 0,61 | 0,57 | 0,45 | 0,33 | 0,27 | 0,48 | 0,52 | 0,45 | 0,98 | 0,99 | 0,99 |
| MedRoBERTa.nl | Confusion | 1600 | 0,61 | 0,54 | 0,56 | 0,40 | 0,33 | 0,35 | 0,46 | 0,29 | 0,34 | 0,98 | 0,99 | 0,99 |
| MedRoBERTa.nl | Cough | 200 | 0,25 | 0,33 | 0,29 | 0,76 | 1,00 | 0,86 | 0,00 | 0,00 | 0,00 | 0,00 | 0,00 | 0,00 |
| MedRoBERTa.nl | Cough | 400 | 0,55 | 0,39 | 0,40 | 0,79 | 0,99 | 0,88 | 0,00 | 0,00 | 0,00 | 0,85 | 0,19 | 0,31 |
| MedRoBERTa.nl | Cough | 600 | 0,59 | 0,59 | 0,59 | 0,93 | 0,96 | 0,95 | 0,00 | 0,00 | 0,00 | 0,85 | 0,81 | 0,83 |
| MedRoBERTa.nl | Cough | 800 | 0,60 | 0,59 | 0,59 | 0,93 | 0,96 | 0,94 | 0,00 | 0,00 | 0,00 | 0,87 | 0,79 | 0,82 |
| MedRoBERTa.nl | Cough | 1000 | 0,60 | 0,60 | 0,60 | 0,94 | 0,96 | 0,95 | 0,00 | 0,00 | 0,00 | 0,87 | 0,85 | 0,86 |
| MedRoBERTa.nl | Cough | 1200 | 0,60 | 0,59 | 0,59 | 0,93 | 0,97 | 0,95 | 0,00 | 0,00 | 0,00 | 0,88 | 0,79 | 0,83 |
| MedRoBERTa.nl | Cough | 1400 | 0,60 | 0,60 | 0,60 | 0,94 | 0,96 | 0,95 | 0,00 | 0,00 | 0,00 | 0,87 | 0,85 | 0,86 |
| MedRoBERTa.nl | Cough | 1600 | 0,60 | 0,59 | 0,59 | 0,93 | 0,97 | 0,95 | 0,00 | 0,00 | 0,00 | 0,88 | 0,79 | 0,83 |
| MedRoBERTa.nl | Crackles upon auscultation | 200 | 0,55 | 0,49 | 0,47 | 0,00 | 0,00 | 0,00 | 0,66 | 1,00 | 0,80 | 0,97 | 0,47 | 0,63 |
| MedRoBERTa.nl | Crackles upon auscultation | 400 | 0,83 | 0,57 | 0,57 | 0,87 | 0,13 | 0,20 | 0,70 | 0,98 | 0,81 | 0,93 | 0,59 | 0,72 |
| MedRoBERTa.nl | Crackles upon auscultation | 600 | 0,88 | 0,77 | 0,81 | 0,81 | 0,77 | 0,79 | 0,85 | 0,94 | 0,89 | 0,97 | 0,61 | 0,74 |
| MedRoBERTa.nl | Crackles upon auscultation | 800 | 0,88 | 0,82 | 0,85 | 0,84 | 0,81 | 0,83 | 0,88 | 0,95 | 0,91 | 0,92 | 0,70 | 0,79 |
| MedRoBERTa.nl | Crackles upon auscultation | 1000 | 0,89 | 0,83 | 0,85 | 0,88 | 0,80 | 0,84 | 0,88 | 0,96 | 0,92 | 0,92 | 0,72 | 0,80 |
| MedRoBERTa.nl | Crackles upon auscultation | 1200 | 0,87 | 0,85 | 0,86 | 0,84 | 0,86 | 0,85 | 0,91 | 0,92 | 0,92 | 0,87 | 0,76 | 0,80 |
| MedRoBERTa.nl | Crackles upon auscultation | 1400 | 0,87 | 0,86 | 0,86 | 0,85 | 0,86 | 0,85 | 0,91 | 0,92 | 0,92 | 0,85 | 0,78 | 0,81 |
| MedRoBERTa.nl | Crackles upon auscultation | 1600 | 0,88 | 0,86 | 0,87 | 0,86 | 0,88 | 0,87 | 0,92 | 0,93 | 0,93 | 0,87 | 0,77 | 0,82 |
| MedRoBERTa.nl | Fever | 200 | 0,48 | 0,37 | 0,29 | 0,54 | 0,03 | 0,06 | 0,46 | 0,20 | 0,23 | 0,45 | 0,88 | 0,59 |
| MedRoBERTa.nl | Fever | 400 | 0,60 | 0,52 | 0,51 | 0,67 | 0,24 | 0,35 | 0,45 | 0,70 | 0,54 | 0,69 | 0,64 | 0,66 |
| MedRoBERTa.nl | Fever | 600 | 0,70 | 0,70 | 0,70 | 0,69 | 0,74 | 0,71 | 0,62 | 0,60 | 0,61 | 0,79 | 0,77 | 0,78 |
| MedRoBERTa.nl | Fever | 800 | 0,73 | 0,73 | 0,72 | 0,68 | 0,79 | 0,73 | 0,74 | 0,54 | 0,62 | 0,77 | 0,85 | 0,81 |
| MedRoBERTa.nl | Fever | 1000 | 0,73 | 0,73 | 0,72 | 0,70 | 0,75 | 0,72 | 0,69 | 0,59 | 0,64 | 0,79 | 0,84 | 0,81 |
| MedRoBERTa.nl | Fever | 1200 | 0,71 | 0,70 | 0,70 | 0,70 | 0,75 | 0,72 | 0,62 | 0,62 | 0,62 | 0,80 | 0,73 | 0,76 |
| MedRoBERTa.nl | Fever | 1400 | 0,72 | 0,72 | 0,72 | 0,70 | 0,81 | 0,74 | 0,67 | 0,61 | 0,64 | 0,80 | 0,75 | 0,77 |
| MedRoBERTa.nl | Fever | 1600 | 0,71 | 0,72 | 0,71 | 0,66 | 0,81 | 0,72 | 0,67 | 0,60 | 0,63 | 0,79 | 0,75 | 0,77 |
| MedRoBERTa.nl | Ill appearance | 200 | 0,23 | 0,33 | 0,27 | 0,00 | 0,00 | 0,00 | 0,00 | 0,00 | 0,00 | 0,69 | 1,00 | 0,82 |

| | | | | | | Per class metrics | | | | | | | | |
|---|---|---|---|---|---|---|---|---|---|---|---|---|---|---|
| | | | Macro-averaged metrics | | | 'Present' | | | 'Absent' | | | 'Not reported' | | |
| Model | Sign or symptom | Number of samples | Precision | Recall | F1-score | Precision | Recall | F1-score | Precision | Recall | F1-score | Precision | Recall | F1-score |
| MedRoBERTa.nl | Ill appearance | 400 | 0,58 | 0,62 | 0,58 | 0,11 | 0,02 | 0,03 | 0,72 | 0,86 | 0,78 | 0,89 | 0,98 | 0,93 |
| MedRoBERTa.nl | Ill appearance | 600 | 0,86 | 0,83 | 0,84 | 0,75 | 0,64 | 0,69 | 0,91 | 0,88 | 0,90 | 0,93 | 0,96 | 0,94 |
| MedRoBERTa.nl | Ill appearance | 800 | 0,87 | 0,81 | 0,83 | 0,78 | 0,57 | 0,64 | 0,91 | 0,89 | 0,90 | 0,92 | 0,96 | 0,94 |
| MedRoBERTa.nl | Ill appearance | 1000 | 0,87 | 0,85 | 0,86 | 0,74 | 0,69 | 0,71 | 0,93 | 0,89 | 0,91 | 0,93 | 0,95 | 0,94 |
| MedRoBERTa.nl | Ill appearance | 1200 | 0,88 | 0,85 | 0,86 | 0,77 | 0,69 | 0,73 | 0,94 | 0,89 | 0,91 | 0,93 | 0,96 | 0,95 |
| MedRoBERTa.nl | Ill appearance | 1400 | 0,90 | 0,85 | 0,87 | 0,81 | 0,70 | 0,75 | 0,94 | 0,89 | 0,92 | 0,94 | 0,97 | 0,95 |
| MedRoBERTa.nl | Ill appearance | 1600 | 0,90 | 0,85 | 0,87 | 0,83 | 0,67 | 0,73 | 0,95 | 0,90 | 0,92 | 0,93 | 0,97 | 0,95 |
| MedRoBERTa.nl | Shortness of breath | 200 | 0,38 | 0,40 | 0,33 | 0,45 | 0,86 | 0,58 | 0,00 | 0,00 | 0,00 | 0,71 | 0,33 | 0,42 |
| MedRoBERTa.nl | Shortness of breath | 400 | 0,43 | 0,48 | 0,44 | 0,53 | 0,84 | 0,65 | 0,00 | 0,00 | 0,00 | 0,77 | 0,60 | 0,66 |
| MedRoBERTa.nl | Shortness of breath | 600 | 0,80 | 0,69 | 0,69 | 0,79 | 0,89 | 0,84 | 0,75 | 0,27 | 0,36 | 0,86 | 0,91 | 0,88 |
| MedRoBERTa.nl | Shortness of breath | 800 | 0,85 | 0,80 | 0,81 | 0,86 | 0,91 | 0,88 | 0,78 | 0,59 | 0,66 | 0,90 | 0,91 | 0,90 |
| MedRoBERTa.nl | Shortness of breath | 1000 | 0,85 | 0,82 | 0,83 | 0,86 | 0,93 | 0,89 | 0,78 | 0,65 | 0,71 | 0,92 | 0,89 | 0,90 |
| MedRoBERTa.nl | Shortness of breath | 1200 | 0,86 | 0,82 | 0,83 | 0,89 | 0,92 | 0,90 | 0,77 | 0,62 | 0,68 | 0,91 | 0,93 | 0,92 |
| MedRoBERTa.nl | Shortness of breath | 1400 | 0,88 | 0,83 | 0,85 | 0,87 | 0,93 | 0,90 | 0,85 | 0,64 | 0,73 | 0,91 | 0,91 | 0,91 |
| MedRoBERTa.nl | Shortness of breath | 1600 | 0,85 | 0,82 | 0,83 | 0,87 | 0,93 | 0,90 | 0,75 | 0,65 | 0,69 | 0,92 | 0,90 | 0,91 |
| MedRoBERTa.nl | Sputum | 200 | 0,24 | 0,33 | 0,28 | 0,00 | 0,00 | 0,00 | 0,00 | 0,00 | 0,00 | 0,73 | 1,00 | 0,85 |
| MedRoBERTa.nl | Sputum | 400 | 0,50 | 0,37 | 0,36 | 0,74 | 0,15 | 0,21 | 0,00 | 0,00 | 0,00 | 0,76 | 0,98 | 0,85 |
| MedRoBERTa.nl | Sputum | 600 | 0,58 | 0,60 | 0,59 | 0,79 | 0,85 | 0,81 | 0,00 | 0,00 | 0,00 | 0,94 | 0,95 | 0,95 |
| MedRoBERTa.nl | Sputum | 800 | 0,58 | 0,62 | 0,60 | 0,78 | 0,90 | 0,84 | 0,00 | 0,00 | 0,00 | 0,96 | 0,95 | 0,95 |
| MedRoBERTa.nl | Sputum | 1000 | 0,80 | 0,70 | 0,71 | 0,80 | 0,89 | 0,84 | 0,64 | 0,25 | 0,33 | 0,96 | 0,95 | 0,95 |
| MedRoBERTa.nl | Sputum | 1200 | 0,83 | 0,77 | 0,79 | 0,84 | 0,87 | 0,86 | 0,68 | 0,49 | 0,56 | 0,95 | 0,95 | 0,95 |
| MedRoBERTa.nl | Sputum | 1400 | 0,83 | 0,79 | 0,80 | 0,82 | 0,88 | 0,85 | 0,71 | 0,56 | 0,61 | 0,96 | 0,94 | 0,95 |
| MedRoBERTa.nl | Sputum | 1600 | 0,81 | 0,78 | 0,78 | 0,86 | 0,87 | 0,86 | 0,63 | 0,51 | 0,54 | 0,95 | 0,95 | 0,95 |
| RobBERT | Chest pain | 200 | 0,28 | 0,33 | 0,31 | 0,00 | 0,00 | 0,00 | 0,00 | 0,00 | 0,00 | 0,85 | 1,00 | 0,92 |
| RobBERT | Chest pain | 400 | 0,28 | 0,33 | 0,31 | 0,00 | 0,00 | 0,00 | 0,00 | 0,00 | 0,00 | 0,85 | 1,00 | 0,92 |
| RobBERT | Chest pain | 600 | 0,56 | 0,51 | 0,49 | 0,54 | 0,58 | 0,53 | 0,20 | 0,01 | 0,03 | 0,92 | 0,94 | 0,93 |
| RobBERT | Chest pain | 800 | 0,57 | 0,57 | 0,54 | 0,61 | 0,57 | 0,52 | 0,16 | 0,20 | 0,16 | 0,93 | 0,94 | 0,94 |

| | | | | | | Per class metrics | | | | | | | | |
|---|---|---|---|---|---|---|---|---|---|---|---|---|---|---|
| | | | Macro-averaged metrics | | | 'Present' | | | 'Absent' | | | 'Not reported' | | |
| Model | Sign or symptom | Number of samples | Precision | Recall | F1-score | Precision | Recall | F1-score | Precision | Recall | F1-score | Precision | Recall | F1-score |
| RobBERT | Chest pain | 1000 | 0,66 | 0,63 | 0,61 | 0,59 | 0,68 | 0,62 | 0,46 | 0,29 | 0,29 | 0,94 | 0,93 | 0,94 |
| RobBERT | Chest pain | 1200 | 0,68 | 0,63 | 0,63 | 0,64 | 0,60 | 0,61 | 0,46 | 0,33 | 0,35 | 0,93 | 0,95 | 0,94 |
| RobBERT | Chest pain | 1400 | 0,72 | 0,60 | 0,60 | 0,58 | 0,70 | 0,62 | 0,65 | 0,16 | 0,24 | 0,94 | 0,95 | 0,94 |
| RobBERT | Chest pain | 1600 | 0,66 | 0,68 | 0,66 | 0,60 | 0,66 | 0,60 | 0,44 | 0,44 | 0,43 | 0,95 | 0,94 | 0,94 |
| RobBERT | Chills | 200 | 0,32 | 0,33 | 0,32 | 0,00 | 0,00 | 0,00 | 0,00 | 0,00 | 0,00 | 0,95 | 1,00 | 0,97 |
| RobBERT | Chills | 400 | 0,32 | 0,33 | 0,32 | 0,00 | 0,00 | 0,00 | 0,00 | 0,00 | 0,00 | 0,95 | 1,00 | 0,97 |
| RobBERT | Chills | 600 | 0,32 | 0,33 | 0,32 | 0,00 | 0,00 | 0,00 | 0,00 | 0,00 | 0,00 | 0,95 | 1,00 | 0,97 |
| RobBERT | Chills | 800 | 0,54 | 0,55 | 0,55 | 0,65 | 0,67 | 0,65 | 0,00 | 0,00 | 0,00 | 0,98 | 0,99 | 0,99 |
| RobBERT | Chills | 1000 | 0,56 | 0,59 | 0,57 | 0,70 | 0,77 | 0,73 | 0,00 | 0,00 | 0,00 | 0,99 | 1,00 | 0,99 |
| RobBERT | Chills | 1200 | 0,55 | 0,58 | 0,56 | 0,65 | 0,76 | 0,70 | 0,00 | 0,00 | 0,00 | 0,99 | 0,99 | 0,99 |
| RobBERT | Chills | 1400 | 0,63 | 0,65 | 0,64 | 0,88 | 0,95 | 0,91 | 0,00 | 0,00 | 0,00 | 1,00 | 1,00 | 1,00 |
| RobBERT | Chills | 1600 | 0,61 | 0,65 | 0,63 | 0,82 | 0,96 | 0,88 | 0,00 | 0,00 | 0,00 | 1,00 | 0,99 | 0,99 |
| RobBERT | Confusion | 200 | 0,32 | 0,33 | 0,33 | 0,00 | 0,00 | 0,00 | 0,00 | 0,00 | 0,00 | 0,97 | 1,00 | 0,98 |
| RobBERT | Confusion | 400 | 0,32 | 0,33 | 0,33 | 0,00 | 0,00 | 0,00 | 0,00 | 0,00 | 0,00 | 0,97 | 1,00 | 0,98 |
| RobBERT | Confusion | 600 | 0,32 | 0,33 | 0,33 | 0,00 | 0,00 | 0,00 | 0,00 | 0,00 | 0,00 | 0,97 | 1,00 | 0,98 |
| RobBERT | Confusion | 800 | 0,36 | 0,34 | 0,34 | 0,00 | 0,00 | 0,00 | 0,10 | 0,02 | 0,03 | 0,97 | 1,00 | 0,98 |
| RobBERT | Confusion | 1000 | 0,34 | 0,35 | 0,35 | 0,03 | 0,04 | 0,03 | 0,02 | 0,03 | 0,02 | 0,97 | 0,99 | 0,98 |
| RobBERT | Confusion | 1200 | 0,39 | 0,37 | 0,37 | 0,20 | 0,10 | 0,13 | 0,00 | 0,00 | 0,00 | 0,97 | 1,00 | 0,98 |
| RobBERT | Confusion | 1400 | 0,38 | 0,39 | 0,38 | 0,00 | 0,00 | 0,00 | 0,17 | 0,19 | 0,17 | 0,97 | 0,99 | 0,98 |
| RobBERT | Confusion | 1600 | 0,48 | 0,45 | 0,46 | 0,20 | 0,20 | 0,20 | 0,27 | 0,17 | 0,21 | 0,97 | 1,00 | 0,98 |
| RobBERT | Cough | 200 | 0,25 | 0,33 | 0,29 | 0,76 | 1,00 | 0,86 | 0,00 | 0,00 | 0,00 | 0,00 | 0,00 | 0,00 |
| RobBERT | Cough | 400 | 0,54 | 0,46 | 0,47 | 0,83 | 0,97 | 0,89 | 0,00 | 0,00 | 0,00 | 0,77 | 0,40 | 0,53 |
| RobBERT | Cough | 600 | 0,60 | 0,58 | 0,58 | 0,92 | 0,97 | 0,94 | 0,00 | 0,00 | 0,00 | 0,89 | 0,75 | 0,81 |
| RobBERT | Cough | 800 | 0,60 | 0,59 | 0,59 | 0,93 | 0,96 | 0,94 | 0,00 | 0,00 | 0,00 | 0,86 | 0,79 | 0,82 |
| RobBERT | Cough | 1000 | 0,61 | 0,60 | 0,60 | 0,94 | 0,97 | 0,95 | 0,00 | 0,00 | 0,00 | 0,88 | 0,83 | 0,85 |
| RobBERT | Cough | 1200 | 0,61 | 0,60 | 0,60 | 0,93 | 0,97 | 0,95 | 0,00 | 0,00 | 0,00 | 0,89 | 0,82 | 0,85 |
| RobBERT | Cough | 1400 | 0,62 | 0,60 | 0,61 | 0,94 | 0,98 | 0,96 | 0,00 | 0,00 | 0,00 | 0,91 | 0,83 | 0,87 |

| | | | Macro-averaged metrics | | | Per class metrics: 'Present' | | | 'Absent' | | | 'Not reported' | | |
|---|---|---|---|---|---|---|---|---|---|---|---|---|---|---|
| Model | Sign or symptom | Number of samples | Precision | Recall | F1-score | Precision | Recall | F1-score | Precision | Recall | F1-score | Precision | Recall | F1-score |
| RobBERT | Cough | 1600 | 0,61 | 0,58 | 0,59 | 0,92 | 0,98 | 0,95 | 0,00 | 0,00 | 0,00 | 0,90 | 0,77 | 0,82 |
| RobBERT | Crackles upon auscultation | 200 | 0,41 | 0,42 | 0,38 | 0,00 | 0,00 | 0,00 | 0,64 | 1,00 | 0,78 | 0,59 | 0,27 | 0,36 |
| RobBERT | Crackles upon auscultation | 400 | 0,87 | 0,71 | 0,76 | 0,89 | 0,56 | 0,67 | 0,79 | 0,97 | 0,87 | 0,93 | 0,61 | 0,73 |
| RobBERT | Crackles upon auscultation | 600 | 0,89 | 0,79 | 0,82 | 0,87 | 0,80 | 0,83 | 0,86 | 0,96 | 0,90 | 0,95 | 0,61 | 0,74 |
| RobBERT | Crackles upon auscultation | 800 | 0,88 | 0,81 | 0,84 | 0,85 | 0,85 | 0,84 | 0,88 | 0,94 | 0,91 | 0,91 | 0,65 | 0,76 |
| RobBERT | Crackles upon auscultation | 1000 | 0,87 | 0,83 | 0,85 | 0,84 | 0,88 | 0,86 | 0,90 | 0,93 | 0,92 | 0,88 | 0,68 | 0,76 |
| RobBERT | Crackles upon auscultation | 1200 | 0,88 | 0,82 | 0,84 | 0,88 | 0,85 | 0,86 | 0,89 | 0,94 | 0,91 | 0,86 | 0,68 | 0,74 |
| RobBERT | Crackles upon auscultation | 1400 | 0,85 | 0,85 | 0,85 | 0,83 | 0,91 | 0,87 | 0,92 | 0,91 | 0,91 | 0,80 | 0,73 | 0,76 |
| RobBERT | Crackles upon auscultation | 1600 | 0,89 | 0,84 | 0,86 | 0,85 | 0,88 | 0,86 | 0,91 | 0,93 | 0,92 | 0,90 | 0,72 | 0,79 |
| RobBERT | Fever | 200 | 0,28 | 0,36 | 0,26 | 0,07 | 0,00 | 0,00 | 0,34 | 0,18 | 0,20 | 0,44 | 0,89 | 0,59 |
| RobBERT | Fever | 400 | 0,54 | 0,46 | 0,42 | 0,48 | 0,08 | 0,13 | 0,41 | 0,84 | 0,55 | 0,72 | 0,48 | 0,57 |
| RobBERT | Fever | 600 | 0,63 | 0,58 | 0,58 | 0,65 | 0,42 | 0,49 | 0,47 | 0,69 | 0,56 | 0,76 | 0,62 | 0,68 |
| RobBERT | Fever | 800 | 0,69 | 0,69 | 0,68 | 0,61 | 0,80 | 0,69 | 0,70 | 0,49 | 0,57 | 0,77 | 0,79 | 0,78 |
| RobBERT | Fever | 1000 | 0,69 | 0,69 | 0,69 | 0,70 | 0,70 | 0,69 | 0,63 | 0,58 | 0,60 | 0,76 | 0,79 | 0,77 |
| RobBERT | Fever | 1200 | 0,72 | 0,69 | 0,68 | 0,73 | 0,66 | 0,66 | 0,63 | 0,64 | 0,61 | 0,80 | 0,77 | 0,77 |
| RobBERT | Fever | 1400 | 0,71 | 0,69 | 0,68 | 0,68 | 0,77 | 0,71 | 0,66 | 0,61 | 0,61 | 0,80 | 0,70 | 0,72 |
| RobBERT | Fever | 1600 | 0,71 | 0,71 | 0,70 | 0,69 | 0,78 | 0,73 | 0,66 | 0,58 | 0,61 | 0,78 | 0,76 | 0,77 |
| RobBERT | Ill appearance | 200 | 0,23 | 0,33 | 0,27 | 0,00 | 0,00 | 0,00 | 0,00 | 0,00 | 0,00 | 0,69 | 1,00 | 0,82 |
| RobBERT | Ill appearance | 400 | 0,74 | 0,64 | 0,61 | 0,63 | 0,10 | 0,16 | 0,68 | 0,86 | 0,76 | 0,90 | 0,94 | 0,92 |
| RobBERT | Ill appearance | 600 | 0,85 | 0,81 | 0,83 | 0,73 | 0,60 | 0,65 | 0,89 | 0,88 | 0,88 | 0,92 | 0,95 | 0,94 |
| RobBERT | Ill appearance | 800 | 0,88 | 0,82 | 0,84 | 0,82 | 0,59 | 0,68 | 0,89 | 0,89 | 0,89 | 0,93 | 0,96 | 0,94 |
| RobBERT | Ill appearance | 1000 | 0,85 | 0,85 | 0,85 | 0,72 | 0,72 | 0,71 | 0,88 | 0,89 | 0,89 | 0,94 | 0,93 | 0,94 |
| RobBERT | Ill appearance | 1200 | 0,89 | 0,84 | 0,85 | 0,81 | 0,65 | 0,72 | 0,91 | 0,89 | 0,90 | 0,93 | 0,96 | 0,95 |
| RobBERT | Ill appearance | 1400 | 0,86 | 0,84 | 0,84 | 0,74 | 0,67 | 0,69 | 0,90 | 0,88 | 0,89 | 0,94 | 0,95 | 0,94 |
| RobBERT | Ill appearance | 1600 | 0,88 | 0,83 | 0,84 | 0,79 | 0,62 | 0,68 | 0,90 | 0,90 | 0,90 | 0,93 | 0,96 | 0,94 |
| RobBERT | Shortness of breath | 200 | 0,39 | 0,41 | 0,36 | 0,46 | 0,85 | 0,59 | 0,00 | 0,00 | 0,00 | 0,71 | 0,37 | 0,47 |
| RobBERT | Shortness of breath | 400 | 0,43 | 0,46 | 0,42 | 0,51 | 0,87 | 0,64 | 0,00 | 0,00 | 0,00 | 0,78 | 0,52 | 0,62 |

| | | | Macro-averaged metrics | | | Per class metrics: 'Present' | | | 'Absent' | | | 'Not reported' | | |
|---|---|---|---|---|---|---|---|---|---|---|---|---|---|---|
| Model | Sign or symptom | Number of samples | Precision | Recall | F1-score | Precision | Recall | F1-score | Precision | Recall | F1-score | Precision | Recall | F1-score |
| RobBERT | Shortness of breath | 600 | 0,76 | 0,60 | 0,57 | 0,74 | 0,86 | 0,79 | 0,73 | 0,05 | 0,09 | 0,82 | 0,88 | 0,84 |
| RobBERT | Shortness of breath | 800 | 0,84 | 0,79 | 0,80 | 0,85 | 0,91 | 0,88 | 0,77 | 0,52 | 0,61 | 0,90 | 0,92 | 0,91 |
| RobBERT | Shortness of breath | 1000 | 0,84 | 0,80 | 0,82 | 0,85 | 0,91 | 0,88 | 0,77 | 0,59 | 0,67 | 0,91 | 0,91 | 0,91 |
| RobBERT | Shortness of breath | 1200 | 0,86 | 0,81 | 0,83 | 0,87 | 0,92 | 0,89 | 0,81 | 0,59 | 0,68 | 0,91 | 0,93 | 0,91 |
| RobBERT | Shortness of breath | 1400 | 0,87 | 0,81 | 0,83 | 0,85 | 0,94 | 0,89 | 0,83 | 0,58 | 0,68 | 0,92 | 0,91 | 0,91 |
| RobBERT | Shortness of breath | 1600 | 0,87 | 0,83 | 0,85 | 0,87 | 0,94 | 0,90 | 0,82 | 0,64 | 0,72 | 0,92 | 0,91 | 0,91 |
| RobBERT | Sputum | 200 | 0,24 | 0,33 | 0,28 | 0,00 | 0,00 | 0,00 | 0,00 | 0,00 | 0,00 | 0,73 | 1,00 | 0,85 |
| RobBERT | Sputum | 400 | 0,54 | 0,48 | 0,49 | 0,78 | 0,46 | 0,56 | 0,00 | 0,00 | 0,00 | 0,83 | 0,97 | 0,89 |
| RobBERT | Sputum | 600 | 0,58 | 0,60 | 0,59 | 0,80 | 0,83 | 0,81 | 0,00 | 0,00 | 0,00 | 0,94 | 0,96 | 0,95 |
| RobBERT | Sputum | 800 | 0,57 | 0,61 | 0,59 | 0,76 | 0,89 | 0,82 | 0,00 | 0,00 | 0,00 | 0,95 | 0,94 | 0,95 |
| RobBERT | Sputum | 1000 | 0,57 | 0,61 | 0,59 | 0,76 | 0,88 | 0,81 | 0,00 | 0,00 | 0,00 | 0,95 | 0,94 | 0,95 |
| RobBERT | Sputum | 1200 | 0,58 | 0,61 | 0,59 | 0,80 | 0,86 | 0,83 | 0,00 | 0,00 | 0,00 | 0,94 | 0,96 | 0,95 |
| RobBERT | Sputum | 1400 | 0,56 | 0,58 | 0,56 | 0,76 | 0,79 | 0,76 | 0,00 | 0,00 | 0,00 | 0,92 | 0,94 | 0,93 |
| RobBERT | Sputum | 1600 | 0,65 | 0,62 | 0,62 | 0,81 | 0,87 | 0,83 | 0,20 | 0,04 | 0,07 | 0,94 | 0,96 | 0,95 |

**Table A4** | The performance of the MedRoBERTa.nl and RobBERT models as prompt-based classifier in terms of recall, precision, and F1-score for different sample sizes. The models were trained to extract a total of nine signs and symptoms related to lower respiratory tract Infection from Dutch primary care electronic health records.

| | | | | | | Per class metrics | | | | | | | | |
|---|---|---|---|---|---|---|---|---|---|---|---|---|---|---|
| | | | Macro-averaged metrics | | | 'Present' | | | 'Absent' | | | 'Not reported' | | |
| **Model** | **Sign or symptom** | **Number of samples** | **Precision** | **Recall** | **F1-score** | **Precision** | **Recall** | **F1-score** | **Precision** | **Recall** | **F1-score** | **Precision** | **Recall** | **F1-score** |
| MedRoBERTa.nl | Chest pain | 1 | 0,01 | 0,33 | 0,03 | 0,00 | 0,00 | 0,00 | 0,04 | 1,00 | 0,08 | 0,00 | 0,00 | 0,00 |
| MedRoBERTa.nl | Chest pain | 2 | 0,01 | 0,33 | 0,03 | 0,00 | 0,00 | 0,00 | 0,04 | 1,00 | 0,08 | 0,00 | 0,00 | 0,00 |
| MedRoBERTa.nl | Chest pain | 3 | 0,01 | 0,33 | 0,03 | 0,00 | 0,00 | 0,00 | 0,04 | 1,00 | 0,08 | 0,00 | 0,00 | 0,00 |
| MedRoBERTa.nl | Chills | 1 | 0,01 | 0,32 | 0,02 | 0,04 | 0,21 | 0,06 | 0,00 | 0,73 | 0,01 | 0,00 | 0,00 | 0,00 |
| MedRoBERTa.nl | Chills | 2 | 0,01 | 0,32 | 0,02 | 0,04 | 0,21 | 0,06 | 0,00 | 0,73 | 0,01 | 0,00 | 0,00 | 0,00 |
| MedRoBERTa.nl | Chills | 3 | 0,01 | 0,32 | 0,02 | 0,04 | 0,21 | 0,06 | 0,00 | 0,73 | 0,01 | 0,00 | 0,00 | 0,00 |
| MedRoBERTa.nl | Confusion | 1 | 0,34 | 0,33 | 0,05 | 0,01 | 0,92 | 0,02 | 0,00 | 0,02 | 0,00 | 1,00 | 0,06 | 0,11 |
| MedRoBERTa.nl | Confusion | 2 | 0,34 | 0,33 | 0,05 | 0,01 | 0,92 | 0,02 | 0,00 | 0,02 | 0,00 | 1,00 | 0,06 | 0,11 |
| MedRoBERTa.nl | Confusion | 3 | 0,34 | 0,33 | 0,05 | 0,01 | 0,92 | 0,02 | 0,00 | 0,02 | 0,00 | 1,00 | 0,06 | 0,11 |
| MedRoBERTa.nl | Cough | 1 | 0,33 | 0,33 | 0,02 | 0,73 | 0,01 | 0,02 | 0,01 | 0,97 | 0,03 | 0,25 | 0,01 | 0,02 |
| MedRoBERTa.nl | Cough | 2 | 0,33 | 0,33 | 0,02 | 0,73 | 0,01 | 0,02 | 0,01 | 0,97 | 0,03 | 0,25 | 0,01 | 0,02 |
| MedRoBERTa.nl | Cough | 3 | 0,33 | 0,33 | 0,02 | 0,73 | 0,01 | 0,02 | 0,01 | 0,97 | 0,03 | 0,25 | 0,01 | 0,02 |
| MedRoBERTa.nl | Crackles upon auscultation | 1 | 0,28 | 0,33 | 0,30 | 0,26 | 0,37 | 0,30 | 0,59 | 0,63 | 0,61 | 0,00 | 0,00 | 0,00 |
| MedRoBERTa.nl | Crackles upon auscultation | 2 | 0,28 | 0,33 | 0,30 | 0,26 | 0,37 | 0,30 | 0,59 | 0,63 | 0,61 | 0,00 | 0,00 | 0,00 |
| MedRoBERTa.nl | Crackles upon auscultation | 3 | 0,28 | 0,33 | 0,30 | 0,26 | 0,37 | 0,30 | 0,59 | 0,63 | 0,61 | 0,00 | 0,00 | 0,00 |
| MedRoBERTa.nl | Fever | 1 | 0,18 | 0,33 | 0,14 | 0,25 | 1,00 | 0,40 | 0,30 | 0,00 | 0,01 | 0,00 | 0,00 | 0,00 |
| MedRoBERTa.nl | Fever | 2 | 0,18 | 0,33 | 0,14 | 0,25 | 1,00 | 0,40 | 0,30 | 0,00 | 0,01 | 0,00 | 0,00 | 0,00 |
| MedRoBERTa.nl | Fever | 3 | 0,18 | 0,33 | 0,14 | 0,25 | 1,00 | 0,40 | 0,30 | 0,00 | 0,01 | 0,00 | 0,00 | 0,00 |
| MedRoBERTa.nl | Ill appearance | 1 | 0,18 | 0,34 | 0,07 | 0,11 | 1,00 | 0,20 | 0,44 | 0,01 | 0,03 | 0,00 | 0,00 | 0,00 |
| MedRoBERTa.nl | Ill appearance | 2 | 0,18 | 0,34 | 0,07 | 0,11 | 1,00 | 0,20 | 0,44 | 0,01 | 0,03 | 0,00 | 0,00 | 0,00 |
| MedRoBERTa.nl | Ill appearance | 3 | 0,15 | 0,34 | 0,07 | 0,11 | 1,00 | 0,20 | 0,34 | 0,01 | 0,03 | 0,00 | 0,00 | 0,00 |
| MedRoBERTa.nl | Shortness of breath | 1 | 0,04 | 0,33 | 0,07 | 0,00 | 0,00 | 0,00 | 0,12 | 1,00 | 0,22 | 0,00 | 0,00 | 0,00 |
| MedRoBERTa.nl | Shortness of breath | 2 | 0,04 | 0,33 | 0,07 | 0,00 | 0,00 | 0,00 | 0,12 | 1,00 | 0,22 | 0,00 | 0,00 | 0,00 |
| MedRoBERTa.nl | Shortness of breath | 3 | 0,04 | 0,33 | 0,07 | 0,00 | 0,00 | 0,00 | 0,12 | 1,00 | 0,22 | 0,00 | 0,00 | 0,00 |
| MedRoBERTa.nl | Sputum | 1 | 0,01 | 0,33 | 0,02 | 0,00 | 0,00 | 0,00 | 0,03 | 1,00 | 0,05 | 0,00 | 0,00 | 0,00 |

| | | | | | | Per class metrics | | | | | | | | |
|---|---|---|---|---|---|---|---|---|---|---|---|---|---|---|
| | | | Macro-averaged metrics | | | 'Present' | | | 'Absent' | | | 'Not reported' | | |
| **Model** | **Sign or symptom** | **Number of samples** | **Precision** | **Recall** | **F1-score** | **Precision** | **Recall** | **F1-score** | **Precision** | **Recall** | **F1-score** | **Precision** | **Recall** | **F1-score** |
| MedRoBERTa.nl | Sputum | 2 | 0,01 | 0,33 | 0,02 | 0,00 | 0,00 | 0,00 | 0,03 | 1,00 | 0,05 | 0,00 | 0,00 | 0,00 |
| MedRoBERTa.nl | Sputum | 3 | 0,01 | 0,33 | 0,02 | 0,00 | 0,00 | 0,00 | 0,03 | 1,00 | 0,05 | 0,00 | 0,00 | 0,00 |
| RobBERT | Chest pain | 1 | 0,01 | 0,33 | 0,03 | 0,00 | 0,00 | 0,00 | 0,04 | 1,00 | 0,08 | 0,00 | 0,00 | 0,00 |
| RobBERT | Chest pain | 2 | 0,01 | 0,33 | 0,03 | 0,00 | 0,00 | 0,00 | 0,04 | 1,00 | 0,08 | 0,00 | 0,00 | 0,00 |
| RobBERT | Chest pain | 3 | 0,01 | 0,33 | 0,03 | 0,00 | 0,00 | 0,00 | 0,04 | 1,00 | 0,08 | 0,00 | 0,00 | 0,00 |
| RobBERT | Chills | 1 | 0,02 | 0,33 | 0,03 | 0,05 | 0,99 | 0,09 | 0,00 | 0,00 | 0,00 | 0,00 | 0,00 | 0,00 |
| RobBERT | Chills | 2 | 0,02 | 0,33 | 0,03 | 0,05 | 0,99 | 0,09 | 0,00 | 0,00 | 0,00 | 0,00 | 0,00 | 0,00 |
| RobBERT | Chills | 3 | 0,02 | 0,33 | 0,03 | 0,05 | 0,99 | 0,09 | 0,00 | 0,00 | 0,00 | 0,00 | 0,00 | 0,00 |
| RobBERT | Confusion | 1 | 0,01 | 0,31 | 0,02 | 0,02 | 0,12 | 0,03 | 0,02 | 0,80 | 0,04 | 0,00 | 0,00 | 0,00 |
| RobBERT | Confusion | 2 | 0,01 | 0,31 | 0,02 | 0,02 | 0,12 | 0,03 | 0,02 | 0,80 | 0,04 | 0,00 | 0,00 | 0,00 |
| RobBERT | Confusion | 3 | 0,01 | 0,31 | 0,02 | 0,02 | 0,12 | 0,03 | 0,02 | 0,80 | 0,04 | 0,00 | 0,00 | 0,00 |
| RobBERT | Cough | 1 | 0,07 | 0,32 | 0,02 | 0,00 | 0,00 | 0,00 | 0,01 | 0,95 | 0,03 | 0,19 | 0,02 | 0,03 |
| RobBERT | Cough | 2 | 0,07 | 0,32 | 0,02 | 0,00 | 0,00 | 0,00 | 0,01 | 0,95 | 0,03 | 0,21 | 0,02 | 0,03 |
| RobBERT | Cough | 3 | 0,07 | 0,32 | 0,02 | 0,00 | 0,00 | 0,00 | 0,01 | 0,95 | 0,03 | 0,21 | 0,02 | 0,03 |
| RobBERT | Crackles upon auscultation | 1 | 0,08 | 0,33 | 0,13 | 0,23 | 1,00 | 0,38 | 0,00 | 0,00 | 0,00 | 0,00 | 0,00 | 0,00 |
| RobBERT | Crackles upon auscultation | 2 | 0,08 | 0,33 | 0,13 | 0,23 | 1,00 | 0,38 | 0,00 | 0,00 | 0,00 | 0,00 | 0,00 | 0,00 |
| RobBERT | Crackles upon auscultation | 3 | 0,08 | 0,33 | 0,13 | 0,23 | 1,00 | 0,38 | 0,00 | 0,00 | 0,00 | 0,00 | 0,00 | 0,00 |
| RobBERT | Fever | 1 | 0,20 | 0,33 | 0,22 | 0,24 | 0,75 | 0,37 | 0,37 | 0,25 | 0,30 | 0,00 | 0,00 | 0,00 |
| RobBERT | Fever | 2 | 0,20 | 0,33 | 0,22 | 0,24 | 0,75 | 0,37 | 0,37 | 0,25 | 0,30 | 0,00 | 0,00 | 0,00 |
| RobBERT | Fever | 3 | 0,21 | 0,33 | 0,22 | 0,24 | 0,75 | 0,37 | 0,37 | 0,25 | 0,30 | 0,00 | 0,00 | 0,00 |
| RobBERT | Ill appearance | 1 | 0,04 | 0,33 | 0,06 | 0,11 | 1,00 | 0,19 | 0,00 | 0,00 | 0,00 | 0,00 | 0,00 | 0,00 |
| RobBERT | Ill appearance | 2 | 0,04 | 0,33 | 0,06 | 0,11 | 1,00 | 0,19 | 0,00 | 0,00 | 0,00 | 0,00 | 0,00 | 0,00 |
| RobBERT | Ill appearance | 3 | 0,04 | 0,33 | 0,06 | 0,11 | 1,00 | 0,19 | 0,00 | 0,00 | 0,00 | 0,00 | 0,00 | 0,00 |
| RobBERT | Shortness of breath | 1 | 0,04 | 0,33 | 0,07 | 0,00 | 0,00 | 0,00 | 0,12 | 1,00 | 0,22 | 0,00 | 0,00 | 0,00 |
| RobBERT | Shortness of breath | 2 | 0,04 | 0,33 | 0,07 | 0,00 | 0,00 | 0,00 | 0,12 | 1,00 | 0,22 | 0,00 | 0,00 | 0,00 |
| RobBERT | Shortness of breath | 3 | 0,04 | 0,33 | 0,07 | 0,00 | 0,00 | 0,00 | 0,12 | 1,00 | 0,22 | 0,00 | 0,00 | 0,00 |
| RobBERT | Sputum | 1 | 0,08 | 0,33 | 0,13 | 0,24 | 1,00 | 0,38 | 0,00 | 0,00 | 0,00 | 0,00 | 0,00 | 0,00 |

| Model | Sign or symptom | Number of samples | Macro-averaged metrics | | | Per class metrics: 'Present' | | | Per class metrics: 'Absent' | | | Per class metrics: 'Not reported' | | |
|---|---|---|---|---|---|---|---|---|---|---|---|---|---|---|
| | | | Precision | Recall | F1-score | Precision | Recall | F1-score | Precision | Recall | F1-score | Precision | Recall | F1-score |
| RobBERT | Sputum | 2 | 0,08 | 0,33 | 0,13 | 0,24 | 1,00 | 0,38 | 0,00 | 0,00 | 0,00 | 0,00 | 0,00 | 0,00 |
| RobBERT | Sputum | 3 | 0,08 | 0,33 | 0,13 | 0,24 | 1,00 | 0,38 | 0,00 | 0,00 | 0,00 | 0,00 | 0,00 | 0,00 |

### C. PROMPTS

**Box C1**. **The original Dutch prompt (and the translated English prompt) that was used for the prompt-based models to extract the signs and symptoms from the Dutch EHRs.**

| **Original (in Dutch)** | **Translated from original (to English)** |
|---|---|
| *“Classificeer de volgende teksten op basis van de aanwezigheid van het symptoom 'hoesten' als volgt:*<br>*Als 'hoesten' wordt vermeld als voorkomend bij de patiënt, label het als 'Aanwezig.'*<br>*Als 'hoesten' wordt vermeld als niet voorkomend bij de patiënt, label het als 'Niet aanwezig.'*<br>*Als 'hoesten' helemaal niet wordt vermeld in de tekst, label het als 'Niet vermeld.'*<br><br>*Voorbeeld(en):*<br>*<insert samples here>*<br><br>*Print exclusief het label bijbehorende aan de volgende tekst:*<br>*<insert to-be-classified text here>”* | *“Classify the following texts based on the presence of the symptom ‘cough’ as follows:*<br>*If ‘cough’ is mentioned as present for the patient, label this as ‘Present’*<br>*If ‘cough’ is mentioned as not present in the patient, label this as ‘Not present’*<br>*If ‘cough’ is not mentioned in the text, label this as ‘Not mentioned’.*<br><br>*Example(s):*<br>*<insert samples here>*<br><br>*Print only the label corresponding to the following text:*<br>*<insert to-be-classified text here>”* |
